# Extracellular Matrix (ECM) Remodeling Marks an Injury-Like Transcriptional State Associated with Poor Survival in Glioblastoma

**DOI:** 10.1101/2025.11.22.25340791

**Authors:** Steven Lehrer, Peter Rheinstein

## Abstract

**Background:** Glioblastoma (GBM) progression is strongly influenced by its microenvironment, yet the contribution of white-matter injury–associated extracellular matrix (ECM) remodeling to human disease remains unclear. Experimental models show that glioma cells induce axonal injury and glial-scar–like ECM responses that accelerate tumor growth.

**Methods:** We curated an ECM Organization Signature reflecting injury-associated matrix remodeling and applied it to bulk RNA-sequencing data from TCGA-GBM. ECM scores were analyzed alongside wound-healing, microglia/TAM activation, and neuronal integrity signatures. Kaplan–Meier curves and age-adjusted Cox models assessed prognostic significance.

**Results:** ECM organization, wound-healing, and microglia/TAM activation formed a coherent injury-response axis inversely correlated with neuronal programs. ECM-high tumors showed significantly different overall survival compared with ECM-low tumors (log-rank *p* = 0.023). In an age-adjusted Cox model, ECM remodeling independently predicted mortality (HR = 1.23, *p* = 0.0156).

**Conclusion:** White-matter injury–associated ECM remodeling is a prominent, clinically meaningful transcriptional state in human GBM and identifies a prognostic microenvironmental subtype.

## Introduction

Glioblastoma (GBM) is the most aggressive primary malignant brain tumor in adults and remains uniformly lethal despite maximal surgical resection, radiotherapy, and temozolomide chemotherapy. Median overall survival remains approximately 12–15 months, a statistic that has changed little over two decades. The major barrier to therapeutic progress is the profound molecular, cellular, and microenvironmental heterogeneity that characterizes GBM, even within a single tumor [1]. While extensive efforts have focused on profiling late-stage surgical specimens, such samples represent only the endpoint of tumor evolution and provide limited insight into the early pathological processes that drive aggressiveness and treatment resistance.

Early interactions between nascent glioma cells and the surrounding brain microenvironment may shape the eventual malignant trajectory of GBM. Recent work highlights a critical and previously underappreciated role for white matter (WM) injury in initiating and accelerating tumor progression. In a comprehensive somatic mouse model study, Clements et al. demonstrated that early glioma cells preferentially infiltrate WM tracts, mechanically injure axons, and induce SARM1-mediated Wallerian degeneration, which in turn accelerates glioma proliferation at distal sites [2]. Wallerian degeneration is the process by which a nerve axon breaks down after an injury, causing the distal part of the axon to disintegrate and be cleared away [3].

Spatial transcriptomics of tumor-bearing WM have revealed enrichment of wound-healing and extracellular matrix (ECM) organization programs, accompanied by reduced myelination and suppressed neuronal activity. These WM injury–associated signatures correlated strongly with tumor density, indicating that early axonal damage is not merely a consequence of infiltration but an active microenvironmental mechanism that fuels malignant growth [4].

This injury-response phenotype is consistent with other studies identifying GBM subpopulations that adopt mesenchymal and ECM-remodeling transcriptional states, including precancerous or early-progenitor-like cells with an “injured-like” phenotype. Such states are enriched for collagen remodeling, fibrotic pathways, cytokine release, microglial recruitment, and glial scar–like remodeling—features reminiscent of CNS injury responses. Collectively, these findings suggest that early GBM evolution incorporates and amplifies endogenous tissue-injury programs, particularly in WM, and that these programs may produce clinically meaningful tumor subtypes [5].

However, despite these mechanistic insights from experimental systems, it remains unclear whether injury-associated ECM remodeling programs are present in human GBM at diagnosis and whether they have clinical prognostic relevance. To address this question, we developed a curated ECM Organization Signature comprising core matrisome and injury-response genes (including *COL1A1, COL3A1, FN1, VCAN, LOX, MMP2, SPARC, TGFBI*) [6]. The matrisome is the complete inventory of all genes encoding extracellular matrix (ECM) proteins and associated factors. In the current study, we applied this signature to bulk RNA-sequencing data from The Cancer Genome Atlas TCGA-GBM cohort and quantified ECM remodeling states across newly diagnosed tumors.

## METHODS — TCGA-GBM Signature Extraction and Analysis

### Data acquisition

Bulk RNA-seq data for TCGA-GBM were downloaded using TCGAbiolinks (GDC data portal). Quantified gene-level RNA-seq counts (“STAR – Counts”) were retrieved along with patient-level clinical annotations. Samples with non-primary tumors or non-RNA-seq data were excluded.

### Expression preprocessing

Raw counts were normalized using edgeR: filtering for expressed genes, computing TMM normalization factors, and transforming counts to log2-counts per million (logCPM). Ensembl gene IDs were mapped to HGNC symbols using biomaRt. Genes mapping to multiple symbols were collapsed by averaging. The final expression matrix was required to contain >90% of signature genes.

### Injury signature scoring

Four WM-injury signatures—ECM organization, wound healing, microglia/TAM activation, and synaptic/neuronal integrity—were derived from mouse white-matter injury experiments [2]. In TCGA-GBM, signature scores were computed using GSVA (ssGSEA-like z-scoring of mean expression) after gene normalization. Gene sets were applied directly to the human orthologs identified during preprocessing. Orthologs are homologous genes in different species, in this case mouse and human, that evolved from a common ancestral gene through speciation [7].

The ECM Organization Signature consists of genes involved in extracellular matrix structure, remodeling, and injury-associated fibrotic responses [6], including collagens (*COL1A1, COL1A2, COL3A1, COL5A1, COL5A2*), adhesion molecules (*FN1, LAMA2, LAMB1, ITGB1*), matrix-modifying enzymes (*MMP2, MMP9, LOX*), and matricellular regulators (*SPARC, TGFBI, VCAN*). The signature score was computed as the mean z-scored expression of these genes and reflects the extent of ECM remodeling and glial-scar–like microenvironmental activation.

Tumor-associated macrophages (TAMs) and activated microglia represent a major cellular component of the glioblastoma microenvironment and play a central role in shaping tumor behavior. In the healthy brain, microglia function as resident immune sentinels, whereas in glioblastoma they become reprogrammed into a pro-tumor, immunosuppressive phenotype characterized by enhanced phagocytic signaling, cytokine release, extracellular matrix remodeling, and support of mesenchymal transition. Infiltrating peripheral macrophages, recruited through a disrupted blood–brain barrier, further expand this myeloid compartment and contribute overlapping transcriptional programs. Because bulk RNA-sequencing does not readily distinguish resident microglia from infiltrating macrophages, these populations are collectively profiled as microglia/TAM activation [8]. High activation scores reflect a glial-scar–like, injury-associated state marked by elevated expression of *AIF1 (IBA1), CD68, CSF1R, C1QA/B/C*, and chemokines that promote tumor invasion and immune evasion. This myeloid program is strongly linked to extracellular matrix remodeling and wound-healing pathways, forming a coordinated injury-response axis that contributes to glioblastoma progression.

### Clinical integration

Sample barcodes were harmonized to 12-character patient IDs. Clinical metadata (age, vital status, days to death/follow-up) were merged by patient ID.

### Statistical analysis

Signature distributions were visualized using histograms. Pairwise Pearson correlations among signatures were calculated using complete-case data and plotted via a heatmap using ggplot2. Negative correlations were interpreted as opposing biological states (e.g., neuronal integrity vs. ECM remodeling). All analyses were performed in R 4.3.

## RESULTS — TCGA-GBM Injury Signature Analysis

To determine whether the transcriptional programs identified in the mouse white-matter (WM) injury model are recapitulated in human glioblastoma, we applied the four core injury signatures—ECM organization, wound-healing/reactive stroma, microglia/TAM activation, and synaptic/neuronal integrity—to bulk RNA-seq data from The Cancer Genome Atlas glioblastoma cohort (TCGA-GBM). All signatures displayed broad, approximately normal distributions across tumors, indicating substantial inter-tumoral heterogeneity in WM-injury–related biology (Figure 1a–d).

**Figure 1.**
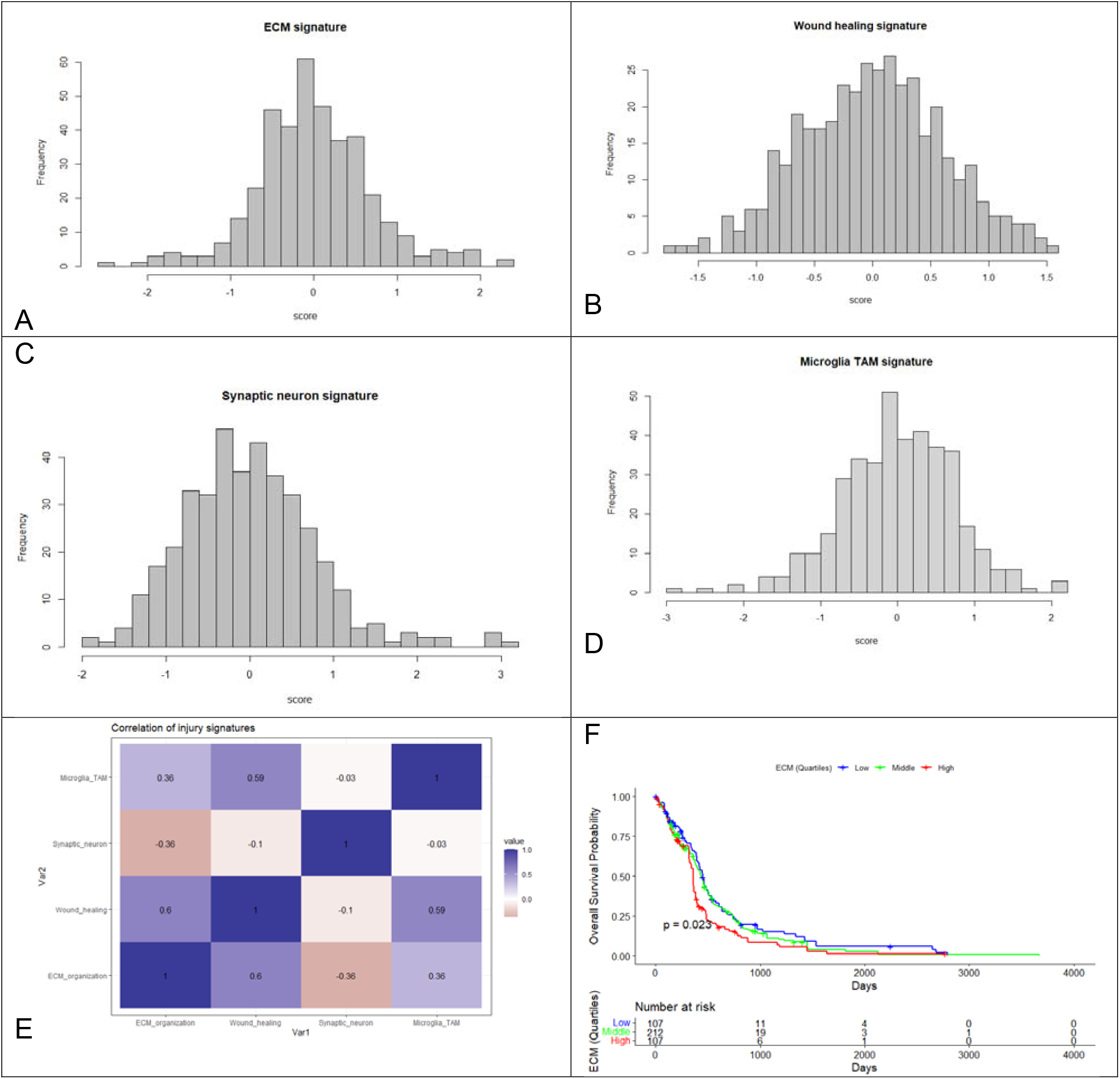
Injury-related transcriptional signatures in TCGA-GBM. (a) Distribution of the ECM organization signature across TCGA-GBM tumors shows a broad, approximately Gaussian spread with a right-tail population exhibiting elevated ECM remodeling. (b) Wound-healing/reactive stroma signature distribution reveals coordinated activation of repair pathways in a subset of tumors. (c) Synaptic/neuronal integrity signature distribution showing reduced neuronal gene expression in tumors with strong injury signatures. (d) Microglia/TAM activation signature, demonstrating a continuum of innate immune activation and macrophage infiltration across tumors. (e) Correlation matrix of the four signatures showing that ECM, wound healing, and TAM activation form a coherent WM-injury axis, while neuronal integrity is inversely correlated with injury signatures. (f) Kaplan–Meier curves comparing overall survival between ECM-high and ECM-low glioblastoma patients in the TCGA-GBM cohort (n = 426). The ECM signature was calculated as the mean z-score of genes involved in extracellular matrix organization and remodeling and dichotomized at the cohort median. Patients with high ECM scores exhibited significantly worse survival compared with ECM-low tumors (log-rank *p* = 0.023). Tick marks indicate censored observations, and the table below the plot shows the number of patients at risk at each time point. Group Median Survival: ECM group Low 448 days, ECM group Middle 432 days, ECM group High 360 days.

The ECM organization signature showed the widest dynamic range, with a distinct right-tail subset of tumors exhibiting heightened matrix remodeling and fibrosis (Figure 1d). This mirrors the transition from healthy WM to reactive glial scar observed in the murine model. The wound-healing signature showed a similar but narrower distribution, consistent with coordinated activation of stromal repair pathways, extracellular matrix deposition, and cytokine signaling.

The microglia/TAM signature revealed a continuum of innate immune activation across GBM, including a prominent right-tail population of tumors enriched for macrophage infiltration and inflammatory cytokines. Notably, the synaptic/neuronal signature was inversely related to the injury signatures: tumors with high ECM and TAM activation showed reduced expression of synaptic and neuronal genes, reflecting loss of intact neural microenvironments as tumors shift toward reactive gliosis and inflammation.

Correlation analysis revealed a coherent injury axis in human GBM. ECM organization strongly correlated with wound-healing (ρ = 0.60) and moderately with TAM activation (ρ = 0.36). Microglia/TAM activation also correlated with wound healing (ρ = 0.59). In contrast, neuronal integrity correlated negatively with both ECM (ρ = 0.36) and wound healing (ρ = 0.10) (Figure 1e). Together, these findings suggest that GBM tumors co-opt WM injury programs—fibrosis, matrix remodeling, and microglial activation—while suppressing neuronal homeostasis, consistent with the glial-scar–like biology observed in the mouse model.

Age-adjusted Cox proportional hazards analysis demonstrated that the ECM organization signature is an independent predictor of poor outcome in TCGA-GBM. After adjusting for age, higher ECM signature scores were associated with a significantly increased hazard of death (HR = 1.23, 95% CI 1.04–1.46, *p* = 0.0156). Age remained a strong covariate, with a 2.2% increase in mortality risk per additional year (*p* = 5.1×10 ^−^ □). The combined model achieved a concordance of 0.61, indicating moderate discriminative ability. These findings support the concept that heightened ECM remodeling reflects a more aggressive tumor microenvironment and contributes independently to GBM progression.

Figure 1(f) shows Kaplan–Meier curves comparing overall survival between ECM-high and ECM-low glioblastoma patients in the TCGA-GBM cohort (n = 426). The ECM signature was calculated as the mean z-score of genes involved in extracellular matrix organization and remodeling and dichotomized at the cohort median. Patients with high ECM scores exhibited significantly worse survival compared with ECM-low tumors (log-rank *p* = 0.023). Group Median Survival: ECM_group_Low 448 days, ECM_group_Middle 432 days, ECM group High 360 days.

These results support a model in which GBM progression involves hijacking endogenous WM-injury responses, creating a permissive stromal and inflammatory microenvironment that resembles chronic glial scar formation. The parallel between mouse injury signatures and human GBM transcriptional states provides strong cross-species validation for the WM-injury hypothesis.

## Discussion

The TCGA-GBM analysis closely mirrors the transcriptional transitions observed in the mouse WM-injury model [2]. In both systems, injury initiates a tripartite response involving:

1. ECM expansion and matrix remodeling
2. Activation of wound-healing and reactive glial pathways
3. Recruitment and activation of microglia/macrophages

Simultaneously, neuronal and synaptic programs decline, reflecting loss of healthy tissue structure and the emergence of a reactive, scar-like niche.

The strong correlations between ECM, wound-healing, and TAM signatures in human GBM demonstrate that tumors adopt an intrinsic injury stromal phenotype like the mouse white-matter response. Conversely, the negative correlation between neuronal and injury signatures indicates that GBM progression involves erosion of neuronal homeostasis and replacement with inflammatory and fibrotic microenvironments.

Together, these findings provide powerful cross-species validation that GBM growth is facilitated by hijacking chronic white-matter injury programs, establishing reactive ECM and immune niches that support invasion, treatment resistance, and neural dysfunction.

Glioblastoma remains one of the most treatment-refractory human cancers, in part because its behavior is shaped as much by its microenvironment as by its intrinsic genetic lesions. Increasing evidence suggests that early interactions between glioma cells and the structural components of white matter (WM) exert profound influence on tumor progression, invasion, and therapy resistance. The emerging WM-injury hypothesis, supported by recent mouse studies and human imaging analyses, proposes that nascent glioma cells actively injure axons, trigger Wallerian-like degeneration, and exploit the resulting inflammatory and fibrotic niche to accelerate malignant expansion [4].

Our study provides new evidence that these injury-associated transcriptional programs are not confined to experimental systems but are a prominent and clinically meaningful feature of human glioblastoma.

By applying WM-injury–derived transcriptional signatures to TCGA-GBM, we identified a coherent “injury axis” composed of extracellular matrix (ECM) organization, wound-healing, and microglia/TAM activation programs. These signatures displayed strong positive correlations with one another, but negative correlations with neuronal/synaptic integrity, indicating that tumors with prominent injury responses tend to suppress neuronal programs and adopt a glial-scar–like phenotype. This transcriptional configuration closely mirrors the state transitions observed in WM infiltrating glioma in mouse models, where axonal damage triggered ECM deposition, microglial recruitment, and reactive stroma formation, each of which contributed to increased tumor density and accelerated growth. The parallels between murine WM injury and human GBM transcriptional states support the view that early microenvironmental disruption is a conserved driver of glioma evolution.

The ECM organization signature emerged as a particularly salient element of this injury axis. Composed of collagens, matrix-modifying enzymes, laminins, fibronectin, and matricellular regulators, the signature reflects the degree of stromal remodeling and fibrotic activation within the tumor microenvironment.

ECM-high tumors demonstrated a markedly distinct clinical phenotype. Although both groups followed the expected steep early decline characteristic of GBM, the ECM-high group exhibited significantly worse overall survival than ECM-low tumors, with clear curve separation and a log-rank p-value of 0.023. In a more stringent age-adjusted Cox model, the ECM signature remained an independent predictor of poor outcome (HR = 1.23, 95% CI 1.04–1.46, p = 0.0156). Together, these results indicate that ECM remodeling captures clinically relevant biology that is not simply a surrogate for patient age or other demographic variables.

The finding that ECM-high tumors have worse prognosis is consistent with a growing literature linking ECM stiffness, matrix crosslinking, collagen deposition, and mechanotransduction to glioma invasiveness and therapeutic resistance. Increased ECM remodeling can enhance integrin signaling, activate focal adhesion kinase (FAK), stiffen the tumor microenvironment, impair T-cell infiltration, and promote mesenchymal transition—all mechanisms associated with more aggressive disease. Moreover, ECM expansion may mimic or amplify aspects of WM injury and glial scar formation, including the recruitment of inflammatory macrophages and the silencing of neuronal activity, thereby creating a feedback loop that reinforces malignant progression. Zhao et al used the SEER database and found that occipital glioblastomas had a distinct survival advantage that may be because of decreased white matter infiltration [9].

Krishna and colleagues have emphasized that gliomas are not passive occupants of the brain but active remodelers of neural circuits, capable of reshaping white-matter structure and function as part of tumor progression. Their work demonstrates that glioma cells co-opt neuronal activity, alter excitatory–inhibitory balance, and engage glial and immune pathways to construct a tumor-permissive microenvironment. Their studies show that glioma infiltration disrupts large-scale connectivity and induces neurophysiologic remodeling, ultimately enabling tumors to integrate into—and exploit— host neural networks. Importantly, this conceptual framework positions GBM not only as a genetic and cellular disease but also as a disorder of tissue-level injury and circuit dysregulation. This aligns closely with emerging evidence from white-matter injury models showing that axonal damage, glial activation, and ECM remodeling are early, conserved responses that gliomas hijack to promote invasion and progression. Incorporating the perspective of Krishna et al. therefore underscores the biological plausibility of GBM adopting an injury-like transcriptional state and supports the relevance of ECM remodeling, microglia/TAM activation, and neuronal loss as key components of a broader CNS injury program exploited by the tumor [10].

Our findings complement mechanistic insights from experimental WM-injury models, which show that glioma-induced axonal damage triggers pro-tumorigenic remodeling of the surrounding stroma. The presence of strong ECM signatures at diagnosis in human tumors suggests that many GBMs may have already undergone extensive microenvironmental injury and remodeling by the time they become clinically detectable. This raises the possibility that early therapeutic intervention targeting ECM-related processes—such as LOX inhibition, integrin blockers, extracellular matrix-modifying enzymes, or agents that preserve axonal integrity—could disrupt the injury-response circuitry that supports tumor expansion.

### Several limitations of our study warrant consideration

Lack of Cellular and Spatial Resolution: The analysis relies on bulk RNA-sequencing (TCGA). This method averages gene expression across the entire tumor sample (tumor cells, microglia, endothelial cells, astrocytes, and ECM). The data cannot definitively identify which cell population is the primary source of the ECM and wound-healing signatures (e.g., is it reactive astrocytes, perivascular cells, or a subset of tumor cells undergoing mesenchymal transition). Future studies using single-cell RNA-seq or spatial transcriptomics are essential to pinpoint the cellular origin of this aggressive phenotype.

Location Confirmation: The conclusion that the signature arises from white matter injury is an inference based on prior literature. The bulk data cannot confirm the spatial location of the gene expression. The ECM activation could be localized primarily to the tumor core or perivascular niche rather than exclusively in infiltrating WM tracts.

Correlation vs. Causality: This is a retrospective correlative study. The high ECM score is a marker of poor survival, but it does not prove that the ECM remodeling causes a poorer outcome. It is possible that the ECM-high state is simply a transcriptional marker for a more aggressive, intrinsically resistant tumor genotype/subtype that is already primed for poor survival.

Trisection of Continuous Data: Dividing the continuous ECM score to create ECM-high, ECM-medium and ECM-low groups for the Kaplan-Meier plot, while visually useful, is an arbitrary cut-off. Presenting the hazard ratio from the continuous score (as we did in the Cox model) is a statistically stronger method, and the conclusions should primarily lean on the Cox model results.

Despite these limitations, our study provides strong evidence that ECM-driven injury programs are a defining feature of human GBM and carry significant prognostic information. The close concordance between mouse WM-injury biology and human tumor transcriptomes strengthens the hypothesis that axonal injury and tissue remodeling are not merely consequences of tumor invasion but may be early, targetable drivers of glioblastoma progression. Therapies aimed at interrupting ECM remodeling, modulating microglial activation, or protecting WM integrity may represent promising avenues for altering the trajectory of this devastating disease.

In summary, our results demonstrate that transcriptional programs associated with white-matter injury—particularly ECM organization and remodeling—are prominently expressed in human glioblastoma and carry significant prognostic value. By bridging mechanistic insights from experimental models with large-scale human tumor transcriptomes, we show that ECM-high tumors exhibit a distinct injury-like microenvironmental state that is associated with worse overall survival, independent of age. These findings support a model in which glioma cells exploit or amplify endogenous CNS injury responses to promote invasion, survival, and therapeutic resistance. Recognizing ECM remodeling as a key axis of GBM progression underscores the need to develop early interventions that target tissue injury, stromal reprogramming, and axon–glia interactions. Such strategies hold potential to disrupt the microenvironmental circuitry that enables GBM growth and may ultimately improve outcomes for patients with this universally lethal disease.

## Data Availability

All data produced in the present work are contained in the manuscript

https://xenabrowser.net/

